# Development of an all-in-one real-time PCR assay for simultaneous detection of spotted fever group rickettsiae, severe fever with thrombocytopenia syndrome virus and orthohantavirus hantanense prevalent in central China

**DOI:** 10.1101/2024.02.26.24303418

**Authors:** Cuixiang Wang, liangjun Chen, xingrong Li, jihong Gu, yating Xiang, Liang Fang, Lili Chen, Yirong Li

## Abstract

Central China has been reported to be one of the most important endemic areas of zoonotic infection by spotted fever group rickettsiae(SFGR), severe fever with thrombocytopenia syndrome virus (SFTSV) and orthohantavirus hantanense(HTNV). Due to similar clinical symptoms, it is challenging to make a definite diagnosis rapidly and accurately in the absence of microbiological tests. In the present study, an all-in-one real-time PCR assay was developed for the simultaneous detection of nucleic acids from SFGR, SFTSV and HTNV. Three linear standard curves for determining SFGR-*ompA*, SFTSV-*L* and HTNV-*L* were obtained within the range of 10^1^-10^6^ copies/μL, with the PCR amplification efficiencies ranging from 93.46% to 96.88% and the regression coefficients R^2^ of >0.99. The detection limit was 1.108 copies/μL for SFGR-*ompA*, 1.075 copies/μL for SFTSV-*L* and 1.006 copies/μL for HTNV-*L*, respectively. Both the within-run and within-laboratory coefficients of variation on the cycle threshold (Ct) values were within the range of 0.53%-2.15%. It was also found there was no statistical difference in the Ct values between with and without other non-target bloodborne virus nucleic acids (P_SFGR-*ompA*_ =0.186, P_SFTSV-*L*_=0.612, P_HTNV-*L*_=0.298). The sensitivity, specificity, positive and negative predictive value were all 100% for determining SFGR-*ompA* and SFTSV-*L*, 97%, 100%, 100% and 99.6% for HTNV-*L*, respectively. Therefore, the all-in-one real-time PCR assay appears to be a reliable, sensitive, rapid, high-throughput and low cost-effective method to diagnose the zoonotic infection by SFGR, SFTSV and HTNV.

**Author Summary:** Spotted fever, severe fever with thrombocytopenia syndrome (SFTS), and hemorrhagic fever with renal syndrome (HFRS) sporadically have outbreaks in central China. Due to the similarities in clinical symptoms and the absence of reliable diagnostic methods, clinical diagnosis and treatment frequently result in misdiagnosis or missed diagnosis. Thus, the development of a fast and accurate diagnostic method is crucial for prevention and precise treatment. In this study, we designed an all-in-one real-time PCR assay to differentiate spotted fever group rickettsiae(SFGR), severe fever with thrombocytopenia syndrome virus (SFTSV) and orthohantavirus hantanense(HTNV). The gene *ompA* of SFGR, as well as the gene segment *L* of SFTSV and HTNV, were used as targets to design primers and probes for amplification. Through the verification of nucleic acid and clinical sample detection, the sensitivity of this detection method exceeded 97%, and its specificity was 100%.

This new assay could be applied in epidemiology and clinical diagnosis, to control new outbreaks, reduce diagnostic and identification time, and improve test efficiency.

## Introduction

Zoonotic infectious diseases typically occur sporadically and are more prevalent in economically underdeveloped areas, such as remote mountainous and forested regions (1-3). Due to the limited medical laboratory resources and similar clinical symptoms, timely and accurate diagnosis of zoonotic infectious diseases is often difficult, which leads to inadequate and untimely treatment. Since 2009, It has been found that there is a high prevalence of severe fever with thrombocytopenia syndrome (SFTS) in central China including Dabie and Yiling Mountains region(4, 5). SFTS is a zoonotic disease infected by a tick-borne virus called severe fever with thrombocytopenia syndrome virus (SFTSV), a novel *Bandavirus* of family *Phenuiviridae*, which was recently named *Dabie Bandavirus* by The International Committee on Taxonomy of Viruses (ICTV). The main clinical manifestations include acute fever, thrombocytopenia, leukopenia, gastrointestinal and neurological symptoms(6-8), moreover, multiple organ failure may occur in severe cases with a maximum mortality of 30% (9, 10). Recently, tick-borne rickettsioses, another zoonotic infectious disease caused by the spotted fever group rickettsiae (SFGR), was found in succession in central China. SFGR is an intracellular bacteria belonging to the spotted fever group (SFG) of the genus *Rickettsia* in the family *Rickettsiaceae* (11). It was reported that the seroprevalence rate of anti-Rickettsia japonica antibody is about 21% among people in Yichang, a city in the Yiling Mountains region of central China(12). The main clinical symptoms of tick-borne rickettsioses also include fever and thrombocytopenia as well as headache, muscle pain, rash and local lymphadenopathy(13-16) (12, 17). It is worth noting that central China including the Dabie and Yiling Mountains region is also known to be an important endemic area for orthohantavirus hantanense (HTNV) infections transmitted by rodents, which belongs to the order *Bunyavirales*, family *Hantaviridae*, and genus *Orthohantavirus*(18). The main epidemic strain of HTNV is HV004 in the past ten years(18-20). Hemorrhagic fever with renal syndrome (HFRS), caused by HTNV, is characterized by a combination of symptoms, which include fever, hemorrhage, thrombocytopenia and acute kidney injury (21, 22). The early clinical manifestations of these pathogens infections are often similar and nonspecific, with most patients experiencing systemic symptoms such as fever, thrombocytopenia, headache, fatigue and muscle aches (23, 24). Therefore, it is challenging to rapidly and accurately identify these pathogens in febrile patients with thrombocytopenia and a history of outdoor activities in central China.

There are a variety of methods including antigen-antibody detection and nucleic acid testing to be used for the identification of microbial pathogens. Antigen-antibody detection such as Enzyme-linked immunosorbent assay and indirect immunofluorescence are limited in the precise diagnosis due to wide antigen cross-reactivity and delayed seroconversion (25-27). Nucleic acid testing has always been considered to be the preferred method for diagnosing viral infections, including quantitative real-time fluorescence PCR assay, isothermal amplification reaction, digital PCR assay and metagenomics next generation sequencing (mNGS). Although isothermal amplification reactions is rapid and do not require a specialized thermocycler, it have high requirements for primer designing and serious challenges for a multiplex PCR assay. Digital PCR assay and mNGS have their advantages in absolute quantification and unbiased microbial infection, respectively, but it is not unsuitable to carry out in hospitals located in economically underdeveloped remote areas due to their high cost or time-consuming procedure. It has been reported that the real-time PCR assay is the preferred choice for detecting the nucleic acid of viruses due to its high accuracy, faster testing time and lower cost. It was also found that there was only one commercially available kit for testing target nucleic acids of SFTSV. Therefore, in order to quickly diagnose these zoonotic infectious diseases caused by SFGR, SFTSV and HTNV in central China, we established a rapid, convenient and accurate all-in-one real-time PCR assay following the evaluation of clinical performances.

## Materials and methods

### Serum or nucleic acid samples

A total of 325 serum or nucleic acid samples were collected in present study. Of them, 17 SFGR DNA-positive nucleic acid samples were obtained from Beijing Center For Disease Control And Prevention (CDC) (n=9) and State Key Laboratory of Virology (n=8), 33 HTNV RNA-positive nucleic acid samples were prepared in State Key Laboratory of Virology (n=21) and Hubei CDC(n=12), the remaining including 46 SFTSV RNA-positive nucleic acid samples and 229 serum samples without three target pathogens were garnered from Zhongnan Hospital of Wuhan University. These nucleic acid samples were previously tested to be positive for SFTSV, Hepatitis B virus (HBV), hepatitis C virus (HCV), Epstein-Barr virus (EBV) and cytomegalovirus (CMV) by commercially available kits (DaAn Gene Co., Ltd), for SFGR DNA and HTNV RNA by Sanger sequencing after nested PCR, respectively(12, 18). The study was approved by the Ethics Committee of Zhongnan Hospital of Wuhan University.

### Primers and probes

All primers and probes were synthesized by General Biosystems (Anhui, China). The detailed primers and probes sequences are listed in **Table 1**. The primers ompA-F/R, and the probe ompA-P were used to amplify the target gene *ompA* in the SFGR. The 5′- ends and 3′-ends of ompA-P were labeled with FAM (6-carboxyfluorescein) and Black Hole Quencher 1 (BHQ1), respectively. The primer pair DL-F/R, and the probe DL-P were employed to amplify the gene segment *L* in SFTSV. The VIC(5-VIC phosphoramidite) and Black Hole Quencher 2 (BHQ2) were labeled in the 5′-ends and 3′-ends of DL-P, respectively. Two sets of primers and probes including HL-F1/R1 and HL-P1, and HL-F2/R2 and HL-P2, were simultaneously adopted to amplify the gene segment *L* in HTNV, and both the 5′-ends and 3′-ends of HL-P1 as well as HL-P2 were labeled with Cy5 (Cyanine 5) and BHQ2, respectively. *Beta-actin (ACTB)* was used as the endogenous reference gene in the all-in-one real-time PCR assay. ACTB-F1, ACTB-R1 and ACTB-P were employed to amplify *ACTB*, and the 5′-ends and 3′-ends of ACTB-P were labeled with 5-Carboxy-X-rhodamine (ROX) and BHQ2. All of the primers and probes for amplifying *ompA* and *L* were designed using PrimerPrimer 5.0 design software, whereas those for amplifying the *ACTB* were the same as those used in previous studies (28). (**Scheme 1**).

**Table 1.**
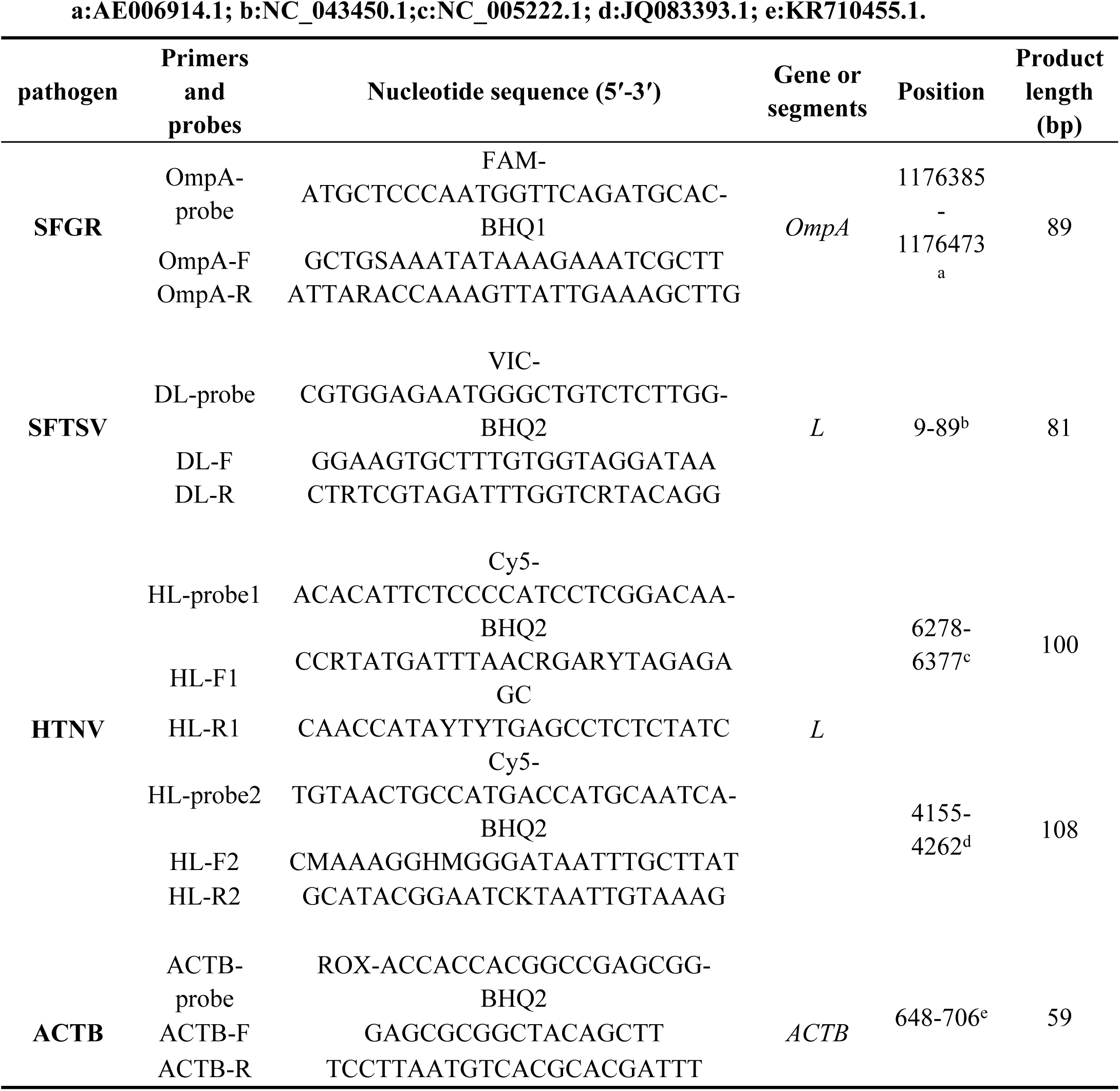
Primers and probes used in the all-in-one real-time PCR assay.

**Scheme 1.**
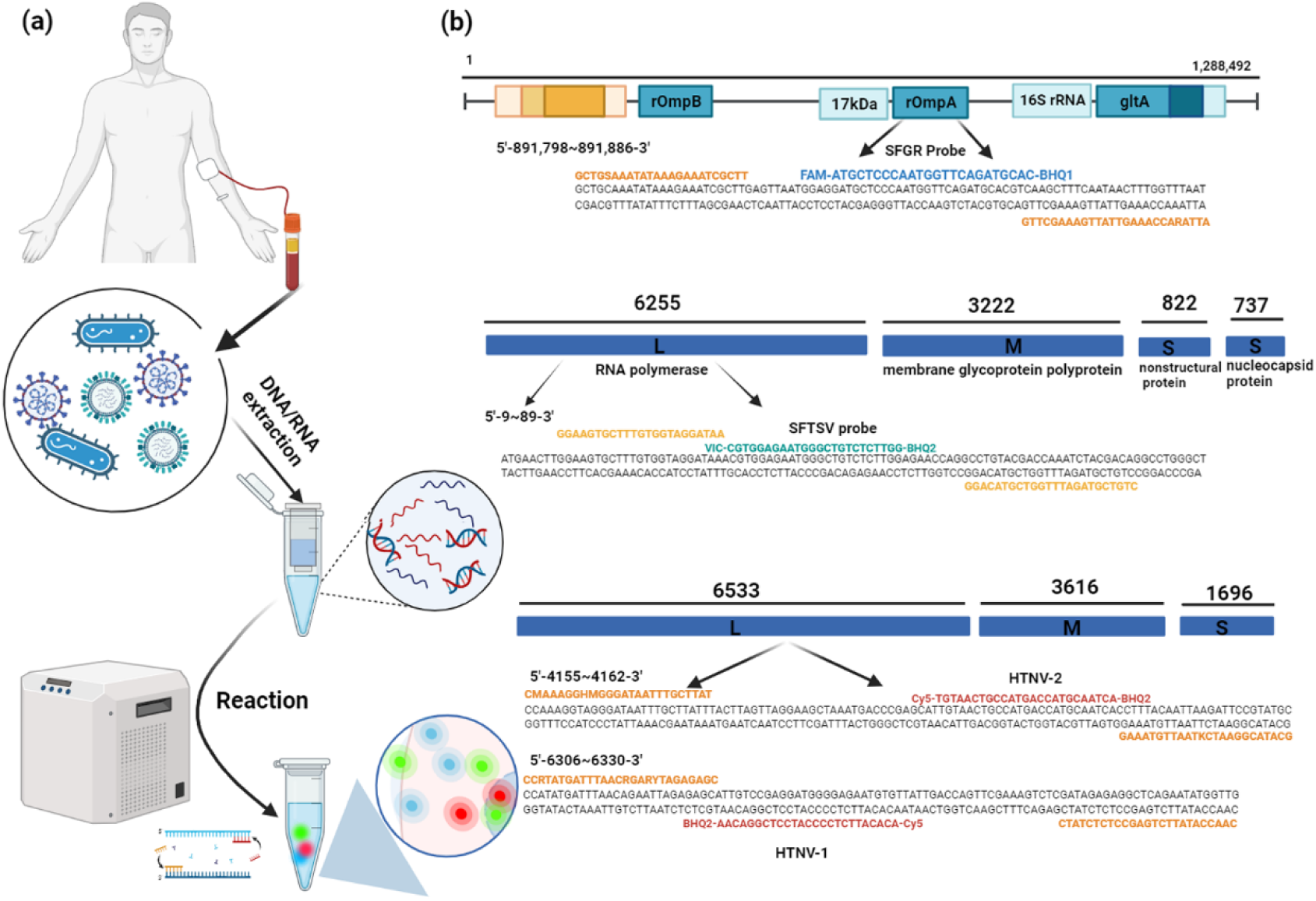
Schematic illustration of the proposed all-in-one real-time PCR assay. (a) Detection principle schematic diagram;(b) Schematic of the SFGR, SFTSV or HTNV genome and corresponding primer sequence design.

### DNA/RNA extraction

DNA and RNA were extracted according to the instructions of the Vazyme Fast Pure Viral DNA/RNA Mini Kit Pro (Vazyme Biotech Co.,Ltd). All of the nucleic acid samples used in the present study were stored at -80 °C until further experiments.

### Construction of the plasmids

Plasmids SFGR-*ompA*, SFTSV-*L*, HTNV-*L*1 and HTNV-*L*2 constructed by General Biol (Anhui) Co., Ltd were used to create standard curves and determine the limit of detection (LOD) for the all-in-one real-time PCR assay. The copy number of plasmid was calculated using the formula: plasmid copy number (copies/μL) = plasmid concentration × 10^-9^ × diluted multiples × 6.02 × 10^23^) / (660 Dalton/bases × DNA length in nucleotides). The initial concentrations of these plasmids were determined to be 6.92 × 10^9^ copies/μL for SFGR-*ompA*, 5.30 × 10^9^ copies/μL for SFTSV-*L*, 6.80 × 10^9^ for HTNV-*L*1 and 5.79 × 10^9^ copies/μL for HTNV-*L*2, respectively. All of them were stored at -80℃ for future experiments.

### All-in-one real-time PCR assay

The all-in-one real-time PCR assay was carried out in an Eppendorf tube for the simultaneous detection of gene *ompA* in the SFGR, segment *L* in both SFTSV and HTNV, and the house-keeping gene *ACTB* with a Gentier 96E/96R real-time thermocycler (Tianlong, Xi’an, China). The final volume of the all-in-one real-time PCR was 25μL, comprising 5.0μL of 5× Neoscript Fast RT Premix Buffer, 1.0μL of 25× Neoscript Fast RTase/UNG Mix, 0.5μL of an ompA-F and ompA-R mixture(300pmol/mL), 0.5μL of a DL-F and DL-R mixture(200pmol/mL), 0.5μL of an HL-F1 and HL-R1 mixture(600pmol/mL), 0.5μL of an HL-F2 and HL-R2 mixture(600pmol/mL), 0.5μL of an ACTB forward and reverse primers mixture. 1μL each of ompA-P, DL-P, HL-P1, HL-P2 and ACTB probes (100pmol/mL), 2μL of template and 9.5μL of ddH2O. The optimized thermal cycling conditions for amplification were as follows: 1 cycle of reverse transcription at 50℃ for 15 minutes and pre-denaturation at 95℃ for 3 minutes, followed by 45 cycles of denaturation at 95℃ for 10 seconds and annealing/ extension at 60℃ for 30 seconds. Monitoring of fluorescence occurred at the extension phase. The Cycle threshold (Ct) values obtained from the all-in-one real-time PCR assay were adopted for the discrimination of the presence of the target gene in clinical samples or not.

### Evaluation of PCR efficiency, LOD and precision of the all-in-one real-time PCR assay

The amplification efficiency of the all-in-one real-time PCR assay was deduced from standard curves as described previously(29), which were generated by plotting the Ct values versus the log_10_ DNA concentration of the standards followed by constructing a linear regression equation. To construct the standard curve, six 10-fold dilutions of the plasmids SFGR-*ompA*, SFTSV-*L*, and HTNV-*L*2 starting with ∼10^6^ copies/μL and ending with ∼10^1^ copies/μL, were yielded, respectively.

LOD was determined according to Chinese National Standard GB/T 37871-2019(30). In brief, plasmids standards at ∼10^1^ copies/μL were absolutely quantified using a digital PCR assay, then were serially two-fold diluted to be about 1 copies/μL. Each diluted plasmid was run in 20 replicates to determine the LOD of the all-in-one real-time PCR assay.

The precision of the all-in-one real-time PCR assay was evaluated according to EP15-A2 (31). In brief, the evaluation was performed per day with three replicate samples at each of two concentrations (∼10^4^ and ∼10^0^ copies/μL) daily for five days. Imprecision was assessed by using the coefficient of variation (CV) on the Ct values.

### Analysis of specificity and interference

A total of 4 nucleic acid samples from other bloodborne viruses were adopted to analyze specificity. Then they were separately tested using the all-in-one real-time PCR assay to investigate nonspecific amplification. In addition, single or multiple target nucleic acids at a concentration of LOD were performed on the all-in-one real-time PCR assay without or with these nucleic acids from other bloodborne viruses in three replicates, then the statistical difference was calculated by an independent samples *t*-test on the Ct values to evaluate interference from HBV, HCV, EBV and CMV nucleic acids.

### Evaluation of the accuracy of the all-in-one real-time PCR assay

To evaluate of accuracy of the all-in-one real-time PCR assay, 321 nucleic acid samples were tested. The results were compared to those reported previously. Sensitivity, specificity, positive and negative predictive values (PPV and NPV) were calculated .

## Results

### Optimal conditions for the all-in-one real-time PCR assay

Our experimental data indicated that the optimal annealing/extension temperature was 60°C (**Table S1**). In addition, to ensure the optimal conditions for detection, the concentrations of primers and probes for amplifying the target gene or gene fragments of SFGR, SFTSV and HTNV were elaboratively optimized. As seen in Fig 1, the optimal concentration of primers ompA-F/R, DL-F/R, HL-F1/R1 and HL-F2/R2 is 300pmol/mL, 200pmol/mL, 600pmol/mL and 600pmol/mL, respectively. While the optimal concentration of all probes is 100pmol/mL. It was shown that when the all-in-one real-time PCR assay had primers and probes at the optimal concentrations, the lowest Ct values were yielded (**Table S2-S4**).

**Fig 1.**
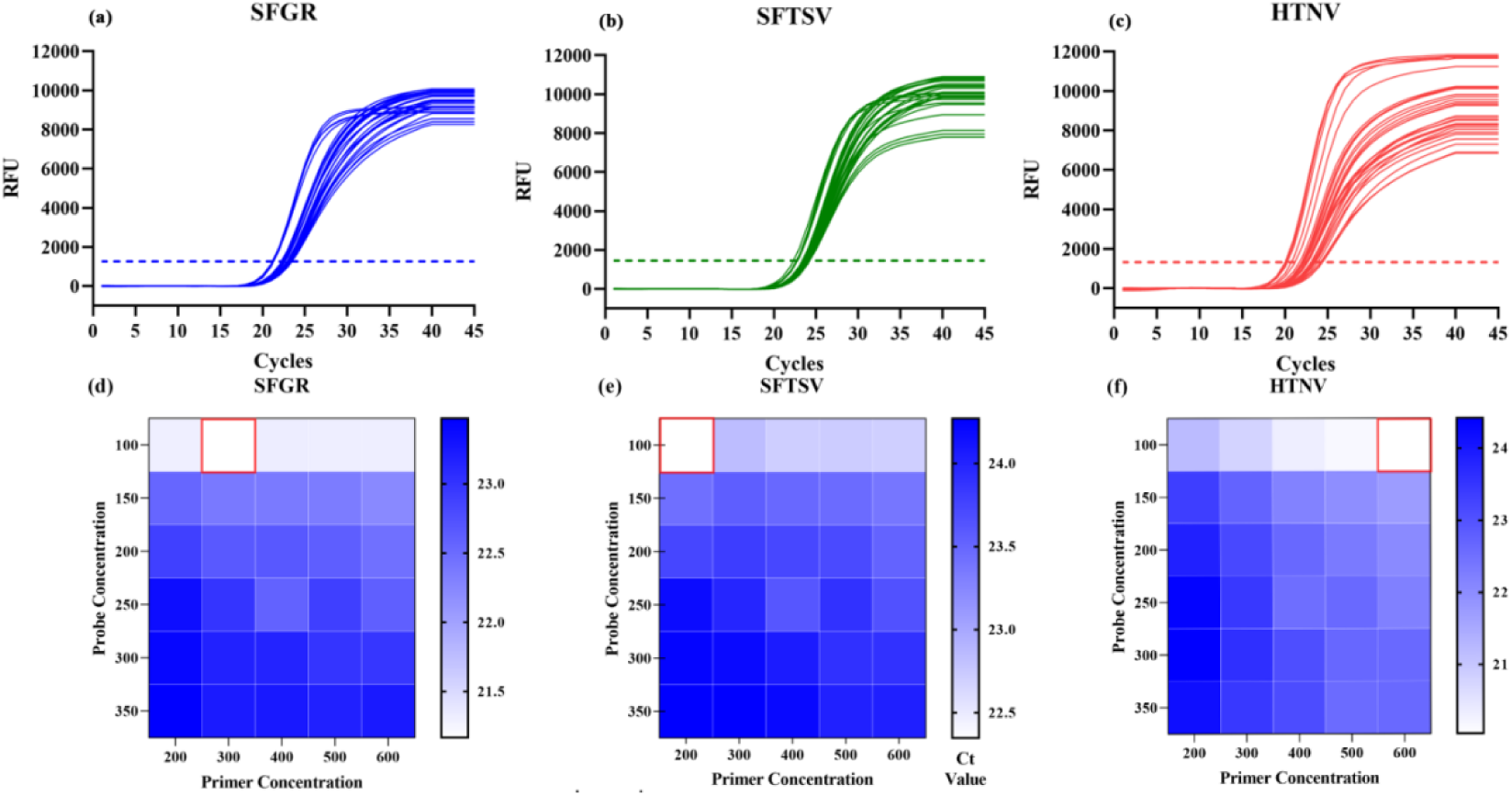
Primer and probe concentration optimization. (a) SFGR’s amplification curve; (b)SFTSV’s amplification curve; (c)HTNV’s amplification curve;(d) SFGR best primer concentration 300pmol/mL, best probe concentration 200pmol/mL; (e) SFTSV best primer concentration 200pmol/mL, best probe concentration 100pmol/mL;(f) HTNV best primer concentration 600pmol/mL, best probe concentration 100pmol/mL

### PCR efficiency, LOD and precision of all-in-one real-time PCR assay

Serial dilutions of four plasmids were co-amplified using the all-in-one real-time PCR assay to construct standard curves. Three linear standard curves were obtained within the range of 10^1^-10^6^ copies/μL with regression coefficients R^2^ ranging from 0.9995 to 0.9999, and amplification efficiencies ranging from 93.46% to 96.88% (**Fig 2**). The LOD were determined to be 1.108 copies/μL for the SFGR-*ompA*, 1.075 copies/μL for the SFTSV-*L* and 1.006 copies/μL for the HTNV-*L* with a detection rate of more than 95%. Regardless of target nucleic acids, the within-run CVs ranged from 0.53%∼1.99%, whereas the within-laboratory CVs were limited to the range between 0.79% and 2.15%. (**Table 2**)

**Fig 2.**
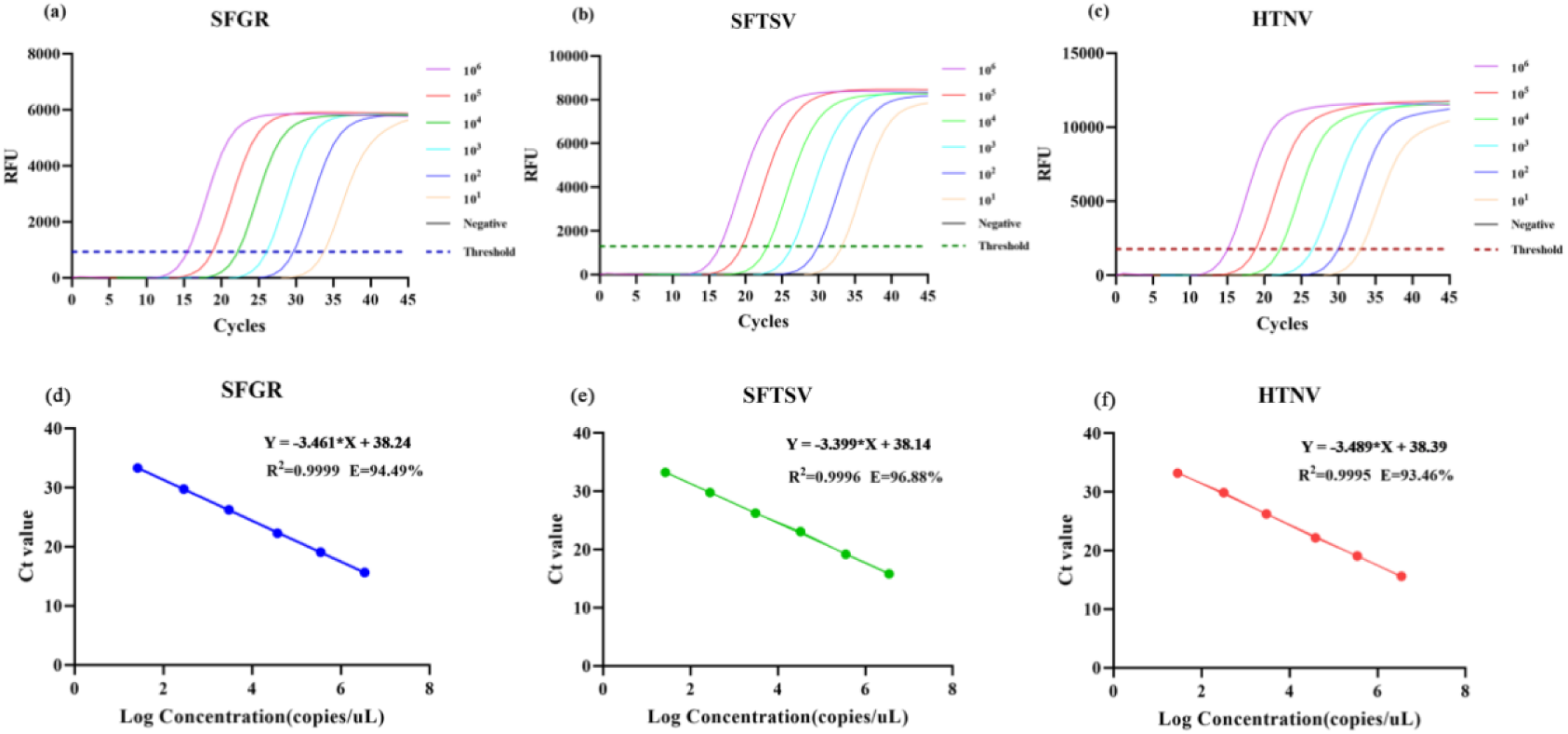
Amplification curves and standard curve construction used about 10^6^-∼10^1^ copies/μL plasmids. (a) SFGR’s amplification curve; (b)SFTSV’s amplification curve; (c)HTNV’s amplification curve; (d) SFGR’s standard curve; (e)SFTSV’s standard curve; (f)HTNV’s standard curve; The all-in-one real-time PCR assay reaction is performed using the optimal primer concentration. 95℃ for 3 minutes; denaturation at 95℃ for 10 seconds; annealing/elongation at 60℃ for 30 seconds. 45 cycles.

**Table 2.** Within-run and within-laboratory reproducibility of the all-in-one real-time PCR assay. SFGR: spotted fever group rickettsiae; SFTSV: severe fever with thrombocytopenia syndrome virus; HTNV: orthohantavirus hantanense.

| Pathogen | Standard concentration (copies/μL) | Within-run | | | Within-laboratory | | |
| --- | --- | --- | --- | --- | --- | --- | --- |
|  |  | Mean Ct | SD | CV(%) | Mean Ct | SD | CV(%) |
| SFGR-*ompA* | 32,629.9 | 20.46200 | 0.10752 | 0.53% | 20.61280 | 0.16201 | 0.79% |
|  | 2.217 | 34.87633 | 0.35874 | 1.03% | 35.17633 | 0.63580 | 1.81% |
| SFTSV-*L* | 36,294.4 | 21.95167 | 0.20802 | 0.95% | 21.83000 | 0.30018 | 1.38% |
|  | 2.151 | 35.80333 | 0.55486 | 1.55% | 35.45907 | 0.68253 | 1.92% |
| HTNV-*L* | 35,303.3 | 20.41267 | 0.14856 | 0.73% | 20.99873 | 0.38912 | 1.85% |
|  | 2.013 | 34.00400 | 0.67554 | 1.99% | 33.94393 | 1.11306 | 2.15% |

### Specificity and the anti-interference ability of the all-in-one **real-time PCR assay**

Four nucleic acid samples which were previously tested positive for HBV-DNA, HCV-RNA, EBV-DNA and CMV-DNA, respectively, were performed to evaluate the specificity of the all-in-one real-time PCR assay. All of them yielded horizontal amplification plots, whereas only plasmids SFGR-*ompA*, SFTSV-*L*, and HTNV-*L* yielded classical “S” type curves (**Fig 3**). Then the plasmids were tested to evaluate the interference from HBV-DNA, HCV-RNA, EBV-DNA and CMV-DNA. The resulting Ct values are listed in Table 3. It was found there was no statistical difference in the Ct values between single fluorescence and multiple fluorescence assays for all three targets (P_SFGR_=0.186, P_SFTSV_=0.612, P_HTNV_=0.298). It was also shown that the Ct value fluctuation was less than 1.1 when the non-target bloodborne virus nucleic acids were added (**Table 3**).

**Fig 3.**
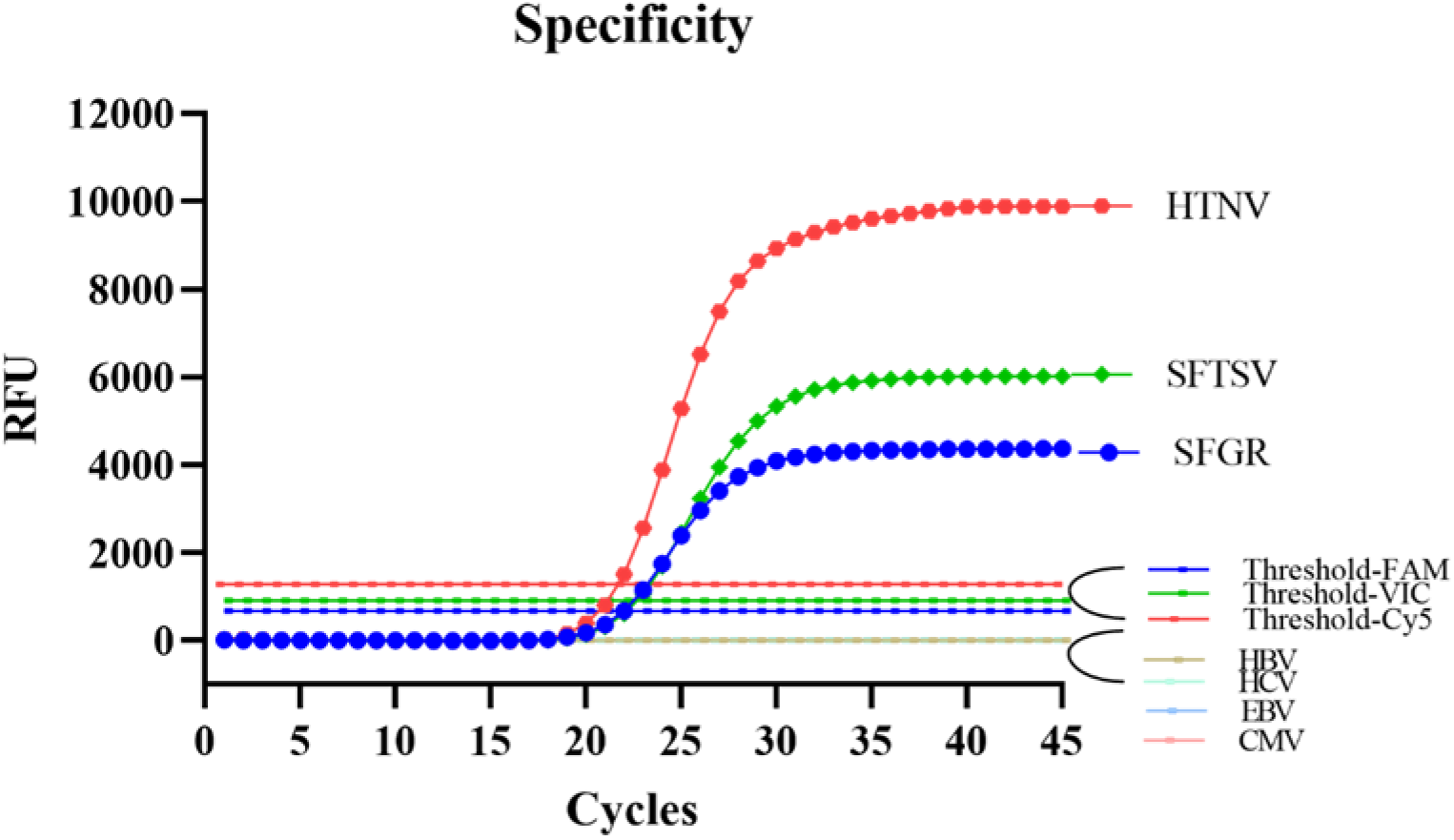
Results of specificity experiment. Only the positive control well has an amplification curve; HBV, HCV, EBV and CMV show no reaction curves.

**Table 3.** The Ct values variation after adding non-specific nucleic acids.

| pathogens |  | Ct values |  |  |  |  |  |  |
| --- | --- | --- | --- | --- | --- | --- | --- | --- |
|  |  |  |  |  | HBV | HCV | EBV | CMV |
| Single template | SFGR-*ompA* | 34.996 | 35.262 | 35.066 | 34.965 | 35.707 | 35.207 | 35.574 |
|  | SFTSV-*L* | 36.926 | 36.645 | 36.520 | 35.918 | 36.121 | 36.121 | 35.887 |
|  | HTNV-*L* | 35.621 | 35.793 | 35.652 | 36.355 | 36.059 | 36.309 | 35.941 |
| Multiple template<sup>a</sup> | SFGR-*ompA* | 35.316 | 35.496 | 35.137 | 35.723 | 35.699 | 36.160 | 35.988 |
|  | SFTSV-*L* | 36.707 | 36.707 | 36.426 | 35.730 | 35.527 | 36.387 | 35.910 |
|  | HTNV-*L* | 35.301 | 35.598 | 35.699 | 36.027 | 36.293 | 35.590 | 36.395 |

<sup>a</sup>Multiple template: SFGR, SFTSV and HTNV mixed as a template (SFGR:1.108 copies/μL; SFTSV:1.075 copies/μL; HTNV:1.006 copies/μL).

### Evaluation accuracy of all-in-one real-time PCR assay with clinical samples

Judgment criteria are as follows: (a) Positive: Ct<37 and a typical amplification curve is observed (Ct_ACTB_22.48-27.67). (b) Negative: no Ct value and no amplification curve(Ct_ACTB_22.48-27.67). (c) Retesting: Ct>37 and a typical amplification curve is observed. If the retest result is the same as mentioned above, it is considered positive; otherwise, it is deemed negative(Ct_ACTB_22.48-27.67). (d) Unqualified DNA sample: Ct_ACTB_<22.48 and Ct_ACTB_>27.67 (Ct_ACTB_ were calculated from the Ct values of 321 characterized samples with mean and standard deviation (SD): 25.07±2.60).

A total of 321 samples were collected to evaluate the accuracy of the all-in-one real-time PCR assay. It was found the sensitivity, specificity, positive and negative predictive value for testing SFGR-*ompA* and SFTSV-*L* nucleic acids were all 100%, whereas the detection of HTNV-*L* was 97%, 100%, 100% and 99.6%, respectively. (**Table 4**)

**Table 4.**
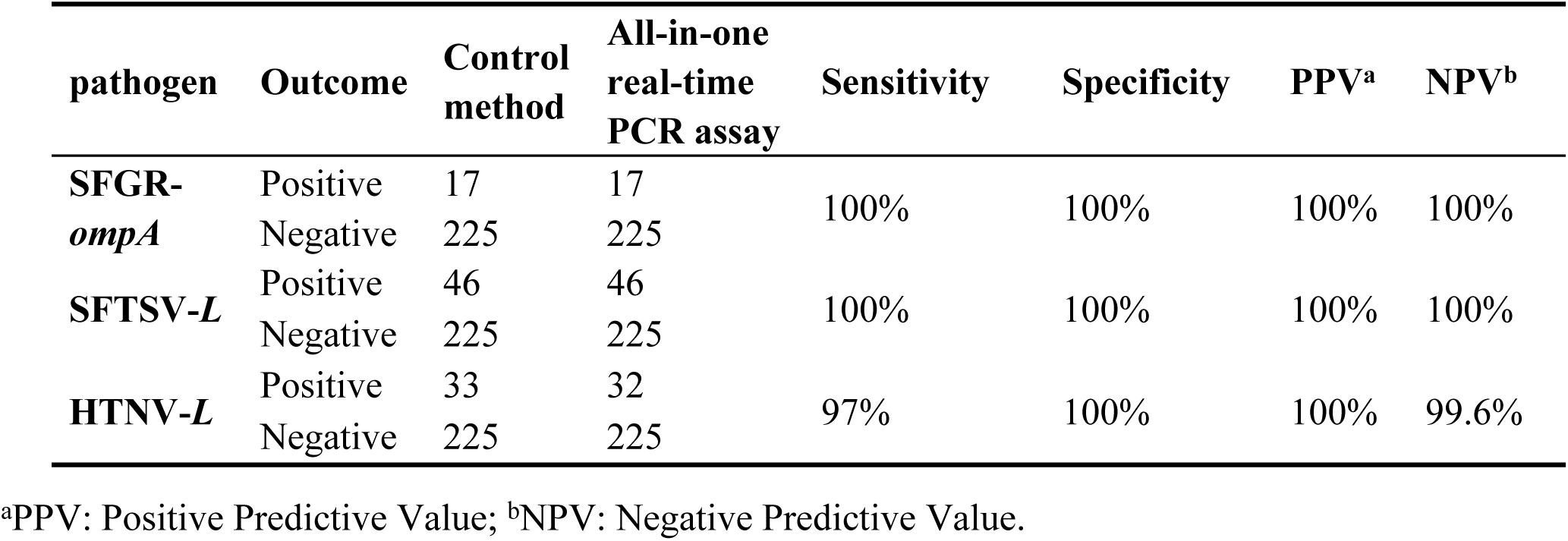
The sensitivity, specificity, PPV and NPV of the all-in-one real-time PCR assay.

| pathogen | Outcome | Control method | All-in-one real-time PCR assay | Sensitivity | Specificity | PPV<sup>a</sup> | NPV<sup>b</sup> |
| --- | --- | --- | --- | --- | --- | --- | --- |
| **SFGR-*ompA*** | Positive | 17 | 17 | 100% | 100% | 100% | 100% |
|  | Negative | 225 | 225 |  |  |  |  |
| **SFTSV-*L*** | Positive | 46 | 46 | 100% | 100% | 100% | 100% |
|  | Negative | 225 | 225 |  |  |  |  |
| **HTNV-*L*** | Positive | 33 | 32 | 97% | 100% | 100% | 99.6% |
|  | Negative | 225 | 225 |  |  |  |  |

<sup>a</sup>PPV: Positive Predictive Value; <sup>b</sup>NPV: Negative Predictive Value.

## Discussion

In the present study, an all-in-one real-time PCR was successfully established to simultaneously detect the nucleic acids from SFGR, SFTSV and HTNV. The SFGR-*ompA*, as well as the gene segment *L* of SFTSV and HTNV, were used as targets for amplification. It was worth noting that two sets of primer and probe were used to amplify gene segment *L* of HTNV due to high variation in China(32). China has had the highest number of HFRS cases worldwide. There exist numerous branches of HTNV, with highly diverse genetics in Heilongjiang, Shanxi, Liaoning, Shandong, Jilin, Hebei, Hunan, Zhejiang, Jiangxi, Jiangsu and Hubei provinces(33). Recently, the genetic evolution analysis of the *L* segment revealed that the viral sequences prevalent in Hubei province cluster together, forming a distinct lineage with genetic variations from viruses in other regions(18). To enhance our detection capabilities, we have designed two sets of primer and probe for HTNV infection. HTNV-*L*2 is primarily utilized to detect the unique lineage (HV004-like) that is prevalent in Hubei province, whereas HTNV-*L*1 is employed for detecting infection in other regions.

The all-in-one real-time PCR method demonstrated high sensitivity, with the ability to detect approximately 1000 copies/ml of the virus genome. It also exhibited an excellent linear range between 10^6^ and 10^1^ copies/μL, where the regression coefficients R^2^ for the target nucleic acids ranged from 0.9995 to 0.9999 and the PCR amplification efficiencies ranged from 93.46% to 96.88%, with a dynamic range of six orders of magnitude (10^1^-^106^ copies/μL). To assess its specificity, we confirmed that other related viruses such as HBV, HCV, EBV and CMV did not produce positive signals and did not affect the Ct value of the positive target, indicating the high specificity of the detection method. Furthermore, the detection method showed high reproducibility, with relatively small variations observed in both intra-assay and inter-assay capabilities. The average coefficients of variation (CVs) for within-run and within-laboratory was limited to the range between 0.53%-2.15%, which is considered acceptable in terms of reproducibility. Using the all-in-one real-time PCR method, we successfully detected SFGR, SFTSV and HTNV in 321 expected clinical samples. Among these samples, 17 were identified as SFGR positive, 46 as SFTSV positive and 33 as HTNV positive. The detection system’s sensitivity, specificity, PPV and NPV for SFGR and SFTSV were all 100%, while the detection of HTNV was 97%, 100%, 100% and 99.6%. These results demonstrate the effectiveness of the all-in-one real-time PCR assay in detecting and differentiating these pathogens in clinical samples.

Since zoonotic infectious diseases like SFTSV, HTNV and SFGR are often prevalent in economically underdeveloped areas, the risk of misdiagnosis is high due to their similar clinical symptoms. Therefore, it is crucial to develop and evaluate a multiplex assay that can detect and identify these similar pathogens simultaneously. So far, there only exists single real-time fluorescence PCR detection methods for SFTSV and HTNV. In comparison, our all-in-one real-time PCR method can simultaneously detect SFGR, SFTSV, and HTNV, making it advantageous for identifying and detecting similar symptoms in patients, as well as conducting large-scale screenings in epidemic areas(34-36). In comparison to the immunochromatographic assay (ICA), which is a cheaper and more convenient on-site testing method but has limitations in terms of sensitivity and delayed detection windows, our method offers a longer detection window and higher sensitivity(37). Compared to the SFTSV CRISPR detection method established by Zou et al., our detection method offers lower detection limits (about 1 copy/μL), reduced costs, and the ability to conduct multiple detections. However, it should be noted that our method has a slightly longer detection time of 60 minutes when compared to their CRISPR detection method(38, 39). For other methods, such as virus isolation, a long experimental cycle of about 10 to 15 days is required in comparison with PCR, thus rendering them unsuitable for clinical promotion. Metagenomic sequencing offers the advantage of detecting a wide range of pathogens without bias and conducting systematic geographical analysis, but its lack of standardization, limited personnel expertise, and high cost hinder its widespread application in clinical practice(40). Overall, the all-in-one real-time PCR assay we have established has the advantages of lower detection limits, lower costs, shorter processing time, and longer detection window period, making it suitable for application in primary medical institutions and remote areas.

Our research demonstrates that all-in-one real-time PCR assay testing has high sensitivity, specificity and reproducibility. Additionally, the turnaround time for experiments is approximately 2 hours, including nucleic acid extraction steps. This makes it a high-throughput, reliable and cost-effective diagnostic and screening tool for early clinical diagnosis of acute-phase SFTSV, HTNV and SFGR. Consequently, the all-in-one real-time PCR assay enables the simultaneous detection of multiple pathogens in a single reaction system, offering great potential for future clinical point-of-care applications. This advancement holds promise in assisting with early and accurate diagnosis, as well as contributing to effective public health management and infectious disease control.

## Data Availability

All relevant data are within the manuscript and its Supporting Information files.

## Acknowledgments

We are grateful to the Beijing Center For Disease Control And Prevention and State Key Laboratory of Virology for providing experimental facilitates.

## Supporting information

**S1 Fig. Digital PCR Results of SFGR 10^3^ copies/μL concentration** (a)The scatter plot of SFGR 10^3^ copies/μL. 3,262.99 copies/μL (b)The scatter plot of SFGR 10^1^ copies/μL. 22.17 copies/μL.

**S2 Fig. Digital PCR Results of SFTSV 10^3^ copies/μL concentration** (a)The scatter plot of SFTSV 10^3^ copies/μL.3,629.44 copies/μL (b)The scatter plot of SFTSV 10^1^ copies/μL.21.51 copies/μL

**S3 Fig. Digital PCR Results of HTNV10^3^ copies/μL concentration** (a)The scatter plot of HTNV 10^3^ copies/μL. 3,530.33 copies/μL (b)The scatter plot of HTNV 10^1^ copies/μL. 20.13 copies/μL

**S1 Table. The Ct value of tempreture optimization.**

**S2 Table. The Ct values correspond to the concentrations of SFGR different gradient primers and probes.**

**S3 Table. The Ct values correspond to the concentrations of SFTSV different gradient primers and probes.**

**S4 Table. The Ct values correspond to the concentrations of HTNV different gradient primers and probes.**

## Uncategorized References

1. Han BA, Kramer AM, Drake JM. Global Patterns of Zoonotic Disease in Mammals. Trends in Parasitology. 2016;32(7):565–77.

2. Plowright RK, Parrish CR, McCallum H, Hudson PJ, Ko AI, Graham AL, et al. Pathways to zoonotic spillover. Nature Reviews Microbiology. 2017;15(8):502–10.

3. Tomori O, Oluwayelu DO. Domestic Animals as Potential Reservoirs of Zoonotic Viral Diseases. Annual Review of Animal Biosciences. 2023;11:33–55.

4. Zhang YZ, He YW, Dai YA, Xiong Y, Zheng H, Zhou DJ, et al. Hemorrhagic fever caused by a novel Bunyavirus in China: pathogenesis and correlates of fatal outcome. Clin Infect Dis. 2012;54(4):527–33.

5. Ren YT, Tian HP, Xu JL, Liu MQ, Cai K, Chen SL, et al. Extensive genetic diversity of severe fever with thrombocytopenia syndrome virus circulating in Hubei Province, China, 2018-2022. PLoS Negl Trop Dis. 2023;17(9):e0011654.

6. Zhuang L, Sun Y, Cui XM, Tang F, Hu JG, Wang LY, et al. Transmission of Severe Fever with Thrombocytopenia Syndrome Virus by Haemaphysalis longicornis Ticks, China. Emerg Infect Dis. 2018;24(5):868–71.

7. Wang M, Huang P, Liu W, Tan W, Chen T, Zeng T, et al. Risk factors of severe fever with thrombocytopenia syndrome combined with central neurological complications: A five-year retrospective case-control study. Front Microbiol. 2022;13:1033946.

8. Li DX. Severe fever with thrombocytopenia syndrome: a newly discovered emerging infectious disease. Clin Microbiol Infect. 2015;21(7):614–20.

9. Li J, Li S, Yang L, Cao P, Lu J. Severe fever with thrombocytopenia syndrome virus: a highly lethal bunyavirus. Crit Rev Microbiol. 2021;47(1):112–25.

10. Yu XJ, Liang MF, Zhang SY, Liu Y, Li JD, Sun YL, et al. Fever with thrombocytopenia associated with a novel bunyavirus in China. N Engl J Med. 2011;364(16):1523–32.

11. Parola P, Paddock CD, Raoult D. Tick-borne rickettsioses around the world: emerging diseases challenging old concepts. Clin Microbiol Rev. 2005;18(4):719–56.

12. Teng Z, Gong P, Wang W, Zhao N, Jin X, Sun X, et al. Clinical Forms of Japanese Spotted Fever from Case-Series Study, Zigui County, Hubei Province, China, 2021. Emerg Infect Dis. 2023;29(1):202–6.

13. Efstratiou A, Karanis G, Karanis P. Tick-Borne Pathogens and Diseases in Greece. Microorganisms. 2021;9(8).

14. Noguchi M, Oshita S, Yamazoe N, Miyazaki M, Takemura YC. Important Clinical Features of Japanese Spotted Fever. Am J Trop Med Hyg. 2018;99(2):466–9.

15. Parola P, Davoust B, Raoult D. Tick- and flea-borne rickettsial emerging zoonoses. Vet Res. 2005;36(3):469–92.

16. Helmick CG, Bernard KW, D’Angelo LJ. Rocky Mountain spotted fever: clinical, laboratory, and epidemiological features of 262 cases. J Infect Dis. 1984;150(4):480–8.

17. Li W, Liu SN. Rickettsia japonica infections in Huanggang, China, in 2021. IDCases. 2021;26:e01309.

18. Chen JT, Zhan JB, Zhu MC, Li KJ, Liu MQ, Hu B, et al. Diversity and genetic characterization of orthohantavirus from small mammals and humans during 2012-2022 in Hubei Province, Central China. Acta Trop. 2023;249:107046.

19. Li JL, Ling JX, Liu DY, Liu J, Liu YY, Wei F, et al. Genetic characterization of a new subtype of Hantaan virus isolated from a hemorrhagic fever with renal syndrome (HFRS) epidemic area in Hubei Province, China. Arch Virol. 2012;157(10):1981–7.

20. Brocato RL, Hooper JW. Progress on the Prevention and Treatment of Hantavirus Disease. Viruses. 2019;11(7).

21. Jonsson CB, Figueiredo LT, Vapalahti O. A global perspective on hantavirus ecology, epidemiology, and disease. Clin Microbiol Rev. 2010;23(2):412–41.

22. Wang ML, Wang JP, Wang TP, Li J, Hui L, Ha XQ. Thrombocytopenia as a Predictor of Severe Acute Kidney Injury in Patients with Hantaan Virus Infections. Plos One. 2013;8(1).

23. Zhao B, Hou HH, Gao R, Tian B, Deng BC. Mononucleosis-like illnesses due to co-infection with severe fever with thrombocytopenia syndrome virus and spotted fever group rickettsia:a case report. Bmc Infectious Diseases. 2021;21(1).

24. Zhan JB, Cheng J, Hu B, Li J, Pan RG, Yang ZH, et al. Pathogens and epidemiologic feature of severe fever with thrombocytopenia syndrome in Hubei province, China. Virus Research. 2017;232:63–8.

25. Lee K, Choi MJ, Cho MH, Choi DO, Bhoo SH. Antibody production and characterization of the nucleoprotein of sever fever with thrombocytopenia syndrome virus (SFTSV) for effective diagnosis of SFTSV. Virol J. 2023;20(1):206.

26. Noden BH, Tshavuka FI, van der Colf BE, Chipare I, Wilkinson R. Exposure and risk factors to coxiella burnetii, spotted fever group and typhus group Rickettsiae, and Bartonella henselae among volunteer blood donors in Namibia. PLoS One. 2014;9(9):e108674.

27. Lederer S, Lattwein E, Hanke M, Sonnenberg K, Stoecker W, Lundkvist Å, et al. Indirect Immunofluorescence Assay for the Simultaneous Detection of Antibodies against Clinically Important Old and New World Hantaviruses. Plos Neglected Tropical Diseases. 2013;7(4).

28. Nakamura A, Nakajima G, Okuyama R, Kuramochi H, Kondoh Y, Kanemura T, et al. Enhancement of 5-fluorouracil-induced cytotoxicity by leucovorin in 5-fluorouracil-resistant gastric cancer cells with upregulated expression of thymidylate synthase. Gastric Cancer. 2014;17(1):188–95.

29. Andersen CB, Holst-Jensen A, Berdal KG, Thorstensen T, Tengs T. Equal performance of TaqMan, MGB, molecular beacon, and SYBR green-based detection assays in detection and quantification of roundup ready soybean. J Agric Food Chem. 2006;54(26):9658–63.

30. China SAotPsRo. Technical specification for quality evaluation of nucleic acid test kit. Standardization Administration of the People’s Republic of China. 2013.

31. R. Neill Carey P, F. Philip Anderson P, Harvey George P, Alfred E. Hartmann M, Verlin K. Janzen M, Anders Kallner M, PhD, et al. <User Verification of Performance for Precision and Trueness;Approved Guideline-Second Edition.pdf>. CLSI. 2006.

32. Liu XJ, Feng JP, Zhang QH, Guo D, Zhang L, Suo T, et al. Analytical comparisons of SARS-COV-2 detection by qRT-PCR and ddPCR with multiple primer/probe sets. Emerging Microbes & Infections. 2020;9(1):1175–9.

33. Zhang S, Wang S, Yin W, Liang M, Li J, Zhang Q, et al. Epidemic characteristics of hemorrhagic fever with renal syndrome in China, 2006-2012. BMC Infect Dis. 2014;14:384.

34. Jiang W, Wang PZ, Yu HT, Zhang Y, Zhao K, Du H, et al. Development of a SYBR Green I based one-step real-time PCR assay for the detection of Hantaan virus. Journal of Virological Methods. 2014;196:145–51.

35. Zeng P, Yang Z, Bakkour S, Wang B, Qing S, Wang J, et al. Development and validation of a real-time reverse transcriptase PCR assay for sensitive detection of SFTSV. J Med Virol. 2017;89(7):1131–8.

36. Jalal S, Hwang SY, Kim CM, Kim DM, Yun NR, Seo JW, et al. Comparison of RT-PCR, RT-nested PCRs, and real-time PCR for diagnosis of severe fever with thrombocytopenia syndrome: a prospective study. Sci Rep. 2021;11(1):16764.

37. Wang XG, Zhang QF, Hao F, Gao XNA, Wu W, Liang MY, et al. Development of a Colloidal Gold Kit for the Diagnosis of Severe Fever with Thrombocytopenia Syndrome Virus Infection. Biomed Research International. 2014;2014.

38. Zuo LL, Miao J, He DM, Fang ZX, Zhang X, Sun CY, et al. Development and characterization of a digital CRISPR/Cas13a based assay for rapid and sensitive diagnosis of severe fever with thrombocytopenia syndrome virus. Sensors and Actuators B-Chemical. 2023;388.

39. Park BJ, Yoo JR, Heo ST, Kim M, Lee KH, Song YJ. A CRISPR-Cas12a-based diagnostic method for multiple genotypes of severe fever with thrombocytopenia syndrome virus. PLoS Negl Trop Dis. 2022;16(8):e0010666.

40. Kim WK, Kim JA, Song DH, Lee D, Kim YC, Lee SY, et al. Phylogeographic analysis of hemorrhagic fever with renal syndrome patients using multiplex PCR-based next generation sequencing. Sci Rep. 2016;6:26017.

